# Cortically Interfaced Human Avatar Enables Remote Volitional Grasp and Shared Discriminative Touch

**DOI:** 10.1101/2025.09.21.25336267

**Authors:** Evan Cater, Sarah Wandelt, Aniket Jangam, Zeev Elias, Erona Ibroci, Christina Maffei, Douglas Griffin, Stephan Bickel, Netanel Ben-Shalom, Adam Stein, Ashesh Mehta, Santosh Chandrasekaran, Chad Bouton

## Abstract

Restoring sensorimotor function in individuals with severe paralysis remains an important challenge in the field of medicine. Here we show, for the first time, an individual with severe upper-limb sensorimotor impairment can interact with real-world objects through another individual using an implanted brain-computer interface (iBCI) and wireless muscle activation, while also engaging in cooperative rehabilitation. This ‘human avatar’ paradigm enabled the iBCI study participant to generate motor intent signals, decoded in real time, to control the hand of other participants wirelessly through high resolution neuromuscular electrical stimulation. Working with an able-bodied participant, the iBCI participant achieved volitional control of the other participant’s hand to discriminate objects of different compliance with 64% accuracy (p < 0.001) under blindfolded conditions. Discrimination was enabled by an S1 encoding paradigm combining multi-electrode recruitment and graded amplitude, which yielded perceptual accuracies exceeding 90% during calibration. Extending this paradigm, the iBCI participant worked with another participant with upper limb impairment and collaborated in a cooperative task: the iBCI participant’s decoded motor commands drove spinal cord and neuromuscular stimulation in the other participant’s arm, enabling bottle pouring with a 94% success rate (p < 0.0001). Qualitative interviews revealed strong subjective satisfaction, highlighting the potential of this form of cooperative rehabilitation. These findings show that cortically interfaced human avatars can restore both motor control and discriminative sensation across individuals, introducing a new framework for cooperative iBCI-based rehabilitation and perhaps even remote human interaction in the future.

## Introduction

Implanted brain-computer interfaces (iBCIs) can provide intention-based control over external devices^1^ and even paralyzed limbs^2,3^. For paralysis applications, an iBCI based ‘neural bypass’ can restore volitional movements in spinal cord injury (SCI) or other neurological conditions^4^. Several studies have demonstrated that users of iBCIs can also modulate their primary motor cortex (M1) neural activity to control virtual hands, control robotic limbs^5^ or maneuver robotic devices^6^ in the real-world. iBCIs have also increasingly incorporated sensory feedback^7–9^, usually through intracortical microstimulation of primary somatosensory cortex (S1), an essential component for achieving natural and precise interactions with the environment^10^. Translating such functional restoration to the user’s own hand^11^, involving both motor control and sensory perception, has been largely unexplored until recently. In a landmark study, the first demonstration of a cortically controlled FES system in humans enabled direct activation of paralyzed muscles after spinal cord injury (SCI) through the user’s thoughts, forming an artificial neural bypass to circumvent the injury entirely^2^. This study, however, did not explore the potential for an *interhuman* neural bypass, where brain-body interface (BBI) technology is employed across two individuals, allowing shared experiences and cooperative tasking.

The word ‘avatar’ comes from the Sanskrit word avatāra, meaning “descent” and has roots in ancient mythology and philosophy where various deities would descend to earth and take on human form. However, in recent years, an avatar often refers to a digital surrogate of oneself. Such digital avatars can be controlled, potentially through a neural interface, to restore^12,13^ or even extend^14^ human capability. A recent study demonstrated the first use of a digital avatar to generate real-time speech and facial expressions using neural activity decoded from the speech motor cortex^15^. Another recent study demonstrated that unidirectional movement control of another individual is possible where participants used EEG-based BCIs to evoke movements in another person via transcranial magnetic stimulation (TMS)^16^. While this approach highlighted the feasibility of interhuman control, it lacked the high-resolution neural recording and decoding necessary for continuous movement control and did not incorporate real-time sensory feedback.

In this work, we demonstrate for the first time, a bidirectional cortically interfaced ‘human avatar’ in which an individual with tetraplegia was able to control the hand movements of other individuals. In one experimental framework, the iBCI participant could control the hand movements of another (able-bodied) person while experiencing ICMS-evoked tactile percepts in real-time. In this sensorimotor human avatar paradigm, the iBCI participant could not only mediate grasping of the human avatar but also could discriminate between multiple objects in the sensorimotor avatar’s hand while blindfolded. In another experimental framework, the iBCI participant performed motor tasks cooperatively with another participant with SCI, helping them significantly increase their performance during real-world object grasping and manipulation. These cortically interfaced, sensorimotor human avatar paradigms represent a significant step in restoring brain-body connection after neurological injury as well as expanding the limits of interhuman communication, interaction, and shared experiences.

## Methods

### Study Participants

The iBCI participant, a male in their 40s, with a high-level (C4-C5) motor and sensory complete SCI with lower motor neuron dysfunction, was implanted with two intracortical electrode arrays in the primary motor cortex (M1) and three electrode arrays in the primary somatosensory cortex (S1). Two additional participants were also enrolled: an able-bodied female in their 20s (able-bodied participant) and a female in their 60s with C5-C7 motor and sensory incomplete SCI (iSCI participant). The study was conducted under a Food and Drug Administration Investigational Device Exemption #G170200 and Northwell Health IRB protocols #17-0840 (NCT03680872) and #20-0425 (NCT04755699), in accordance with the Declaration of Helsinki. All participants provided written informed consent.

### Surgical Implantation

A more detailed description of pre-surgical planning and surgical implantation of five microelectrode arrays into the hand knob gyrus of left primary motor cortex (M1) and hand area of left primary somatosensory cortex (S1) is described elsewhere^11^. Briefly, motor and sensory tasks of the hand performed during fMRI acquisition generated activation maps that guided M1 and S1 array implantation. Intraoperative cortical surface stimulation further identified the precise locations for implantation. The iBCI participant was implanted with five 10×10, 1.5 mm platinum microelectrode arrays with sputtered iridium oxide film (SIROF) coated tips (Neuroport Array, Blackrock Neurotech) in the hand areas of M1 (2×64-channel arrays, 128 total) and S1 (3 sparsely populated arrays, 32-channel each; 96 total) in the left hemisphere.

### S1 Stimulation Discrimination: Paradigm Selection

For providing tactile feedback, intracortical microstimulation was delivered via CereStim multichannel microstimulation system (Blackrock Neurotech, Salt Lake City, Utah) to S1 electrodes. Electrodes which elicited sensation in the iBCI participant’s right distal index finger (distal D2) were identified (Figure 1c). Eight stimulation paradigms were tested to assess the iBCI participant’s accuracy in discriminating three levels of stimulation intensity. The paradigms differed based on which parameters were varied such as amplitude, frequency, ramping of amplitude or frequency, number of electrodes, and combinations therein, as described in Table 1. We also chose to explore multielectrode stimulation to allow for a greater variation in total stimulation, as well as an added dimension of spatial distribution of perceived signal. During the learning phase of each stimulation paradigm, stimulus trains designed with three predetermined levels of the variable parameter of choice (signifying low, medium, or high percept intensities) were delivered to S1, while an investigator announced the intensity level. Each level was repeated three times, totaling nine presentations in random order. Following this, in the discrimination phase, 30 consecutive stimulations were presented (10 of each level, randomized), with 10 seconds between stimulations. After each stimulation, the iBCI participant announced the perceived intensity level. All stimulations were 2 seconds in duration, with a pulse width of 300 μs.

**Table 1:** S1 Encoding Paradigms.

| **Paradigm** | **Low Stimulation** | **Medium Stimulation** | **High Stimulation** | **Performance** | **P-value** |
| --- | --- | --- | --- | --- | --- |
| **Paradigm 1: Amplitude**<br>(100 Hz, Electrode #89) | 40 μA | 70 μA | 100 μA | 56.7% | 0.0059 |
| **Paradigm 2: Frequency**<br>(70 μA, Electrode #89) | 50 Hz | 100 Hz | 200 Hz | 60.0% | 0.0018 |
| **Paradigm 3: Amplitude + Frequency**<br>(Electrode #89) | 40 μA + 50 Hz | 70 μA + 100 Hz | 100 μA + 200 Hz | 80.0% | < 0.001 |
| **Paradigm 4: Amplitude Ramping**<br>(2 seconds stimulation, 100 Hz, Electrode #89) | Linear ramping (40-100 μA) from 0-1.5 sec hold max for 0.5 sec | Linear ramping (40-100 μA) from 0-1.0 sec hold max for 1.0 sec | Linear ramping (40-100 μA) from 0-0.5 sec, hold max for 1.5 sec | 56.7% | 0.0059 |
| **Paradigm 5: Frequency Ramping**<br>(2 seconds stimulation, 70 μA, Electrode #89) | Linear ramping (20-200 μA) from 0-1.5 sec, hold max for 0.5 sec | Linear ramping (20-200 μA) from 0-1.0 sec, hold max for 1.0 sec | Linear ramping (20-200 μA) from 0-0.5 sec, hold max for 1.5 sec | 53.3% | 0.0166 |
| **Paradigm 6: Amplitude + Frequency Ramping**<br>(Electrode #89) | Linear ramping (40-100 μA and 20-200 Hz) from 0-1.5 sec hold max for 0.5 sec | Linear ramping (40-100 μA and 20-200 Hz) from 0-1.0 sec hold max for 1.0 sec | Linear ramping (40-100 μA and 20-200 Hz) from 0-0.5 sec, hold max for 1.5 sec | 53.3% | 0.0166 |
| **Paradigm 7: Electrode Number**<br>(100 μA, 50 Hz) | 1 Electrode (#89) | 5 Electrodes (#65, 68, 74, 81, 86) | 9 Electrodes (#65, 66, 68, 73, 74, 81, 82, 86, 89) | 90% | < 0.001 |
| **Paradigm 8:**<br>**Electrode Number**<br>**+ Amplitude**<br>**(50 Hz)** | 1 Electrode<br>(#89) at 40<br>μA | 5 Electrodes<br>(#65, 68, 74,<br>81, 86) at 70<br>μA | 9 Electrodes<br>(#65, 66, 68,<br>73, 74, 81,<br>82, 86, 89) at<br>100 μA | 93.3% | < 0.001 |

**Fig 1.**
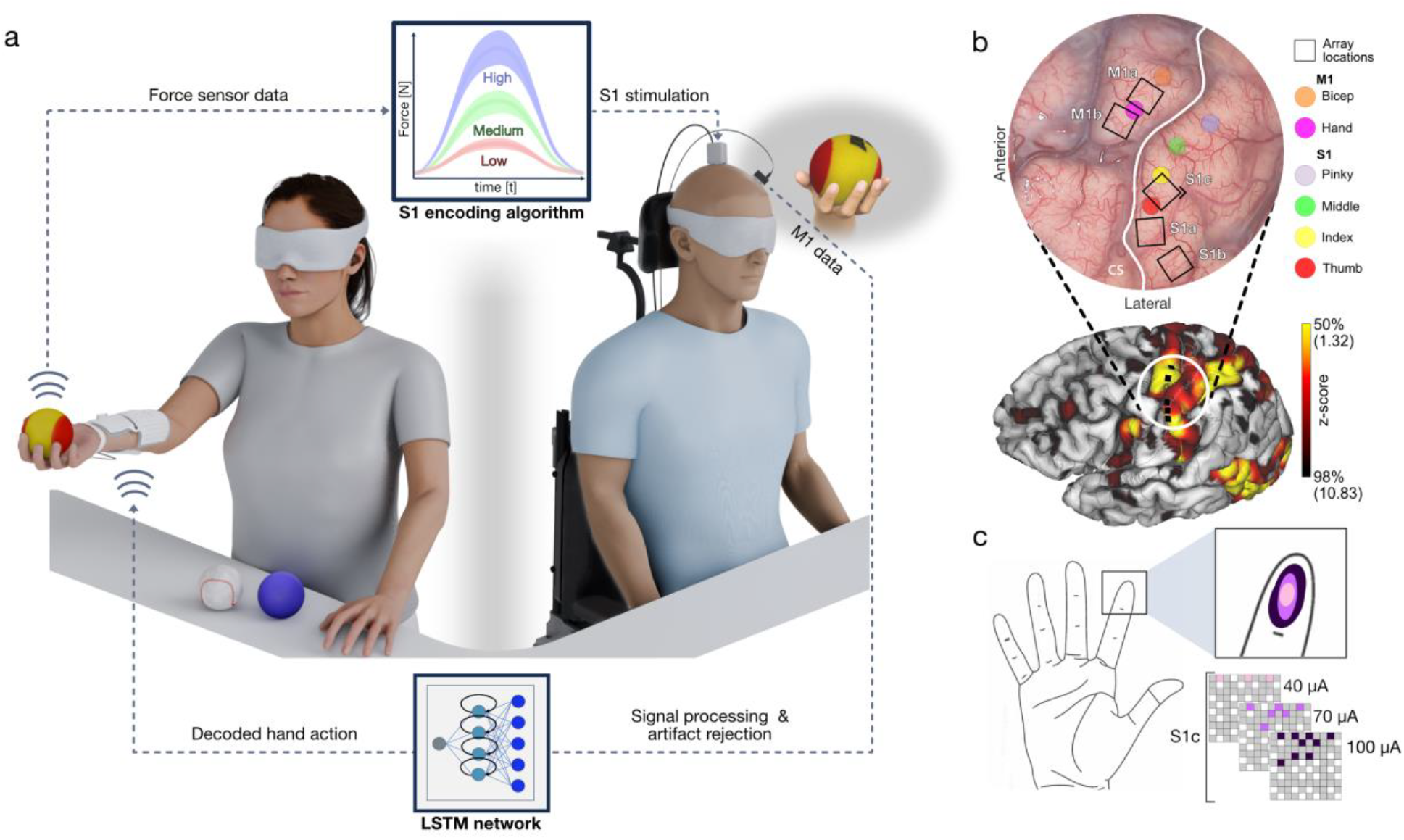
Human avatar experimental paradigm for object grasping and sensory discrimination. **a)** Experimental setup, including iBCI participant (right) with intracortical microelectrode arrays in M1 and S1 and able-bodied participant (left) grasping an object. M1 data is processed via an LSTM neural network to decode grasp intention, prompting NMES-mediated closing of able-bodied participant’s hand around one of three objects. Data collected from force sensors during grasp is transformed via the S1 stimulation profile encoding algorithm to determine S1 stimulation parameters. **b)** Intraoperative image of the brain showing M1, S1 and central sulcus (white). Squares show locations of array implantation. Colored circles show sites where intraoperative cortical stimulation sites evoked EMG activation (M1) or sensory percepts (S1). fMRI response to video-guided and attempted thumb flexion and extension **c)** Map of microelectrode array S1c and corresponding percepts per subset of electrodes as reported by iBCI participant, localized to D2 tip. Pink = subset of 3 electrodes at 40 μA, light purple = 5 electrodes at 70 μA, dark purple = 9 electrodes at 100 μA.

The iBCI participant was asked about the area of percept elicited by the three intensity levels of the highest-performing paradigm. Because the iBCI participant lacked residual movement, experimenters drew the described area on a hand diagram and then further adjusted the drawn area with rounds of feedback from the iBCI participant until the drawn area accurately represented the percept (Figure 1c). The paradigm with the highest accuracy rate informed the parameters of the encoding algorithm described below.

### Object Discrimination Task

A rendering of the experimental set up is shown in Figure 1a. Three objects of similar size and shape but different compliances were chosen – a soft foam ball, a dense foam ball, and a baseball. A custom, wireless high resolution NMES device with 104 embedded electrodes controlled by an on-board microprocessor was attached with flexible straps to the volar aspect of the forearm of the able-bodied participant. Initially, the set of NMES electrodes and the stimulation amplitude optimized for evoking a strong grasp involving all five digits were identified manually and saved to a grasp profile. This profile was modified as needed to trigger adequate grasping before every session.

Two force sensors (Honeywell FMA, 0-5 Newtons range) were used to record the force generated at the distal pads of the thumb (D1) and index finger (D2) of the able-bodied participant’s hand while grasping the objects. The sensors were connected to an ESP32 feather microcontroller, sampled at 10 Hz and transmitted the data via Bluetooth. The force data recorded from these sensors were used to select the precise stimulation parameters for dynamic stimulation of S1 microelectrodes via CereStim R96 (Blackrock Neurotech, Salt Lake City, UT).

A learning phase of 15 object presentations (5 presentations of each object in random order) preceded a discrimination phase which had 30 object presentations (10 per object) for computer-mediated trials, or 21 object presentations (7 per object) for cortically mediated trials. During the learning phase, the iBCI participant was provided visual and auditory feedback in addition to S1 stimulation, i.e., the object was grasped by the able-bodied participant in view of the iBCI participant, and the object identity was announced verbally by investigators. During the discrimination phase, the able-bodied participant was out of view of the iBCI participant, and no verbal indication of object identity was provided. During both learning and discrimination phases, the able-bodied participant was blindfolded.

### Computer-Mediated Trials

During computer-mediated trials, NMES-mediated grasping of the able-bodied participant’s hand was linked to a 6 second visual cue of a virtual hand grasping and opening on a monitor. The NMES pattern was a 3 second exponential ramp up to maximum amplitude (V_out_= V_max_ × (1 – e^(-t/τ)^)), a 2 second hold at maximum amplitude, and a 1 second linear ramp down to 0. There was a 10 second rest interval between the cues.

### Cortically Mediated Trials

Cortically mediated trials utilized a ‘free play’ paradigm, in which the iBCI participant was blindfolded and attempted to initiate a grasping action in the able-bodied participant’s hand. The objects were placed on the able-bodied participant’s hand for 15 seconds each. If the iBCI participant initiated the grasp and identified an object in less than 15 seconds, the next object presentation was initiated without waiting for the full 15 seconds to elapse. When M1 decoding resulted in a grasp signal, NMES grasping ramped up to maximum amplitude over 2 seconds and was sustained by continued successful decoding of a grasp signal from M1 data. When a grasp signal was no longer decoded, NMES grasping returned to rest until the next decoded grasp. Two cortically mediated trials were performed during one session.

### Encoding Algorithm

During the object discrimination task, object contact forces were recorded in real-time by the force sensors placed on the thumb and index finger of the able-bodied participant during grasping. The forces recorded by both sensors were summed to get the total force (*F*). The stimulation amplitude I was determined by F using a piecewise function –

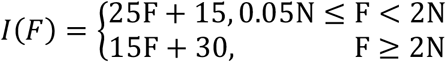

The number of electrodes (n) was determined by the magnitude of F,

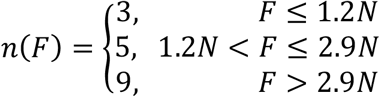

This algorithm was determined with the goals of maximizing the differences between stimulation amplitude for the three objects while remaining under the maximum of 100 μA, and stimulating via 3, 5, and 9 electrodes for the soft foam, dense foam, and baseball, respectively. To mitigate drift in the force values, a running mean of 1.5 seconds was implemented. The running mean was implemented only when NMES grasping was not occurring. The stimulation frequency and pulse width were kept constant at 50 Hz and 300 μs respectively.

### Data Collection and Experimental Setup

Training data for the decoder was obtained on the same day as the experiment in 2 blocks of 15 trials each. The iBCI participant was instructed via screen prompts to alternate between closing their hand for 4 seconds and resting for 4 seconds. The first half was recorded without S1 stimulation, and the second half was recorded with continuous S1 stimulation. This allowed us to incorporate neural changes in M1 due to S1 stimulation in the dataset. Neural signals were acquired at 10 kHz and artifact removal was performed by implementing hardware-triggered software blanking of a total of 12 ms for each stimulation pulse. To prevent discontinuities, while preserving power modulation features, a first derivative was performed followed by zeroing abnormally high signals (high derivatives in original signal indicative of residual artifact) and then transforming to the frequency domain using a short-time Fourier transform (STFT) with a non-overlapping 100 ms Blackman window. Amplitude information was then integrated (summed) across 100-5000 Hz and smoothed using a 1-second boxcar filter. Base 10 log compression was then performed, drift was removed by performing a 24-second running mean subtraction from each signal, and the final features were multiplied by 20 (converting to dB).

### Feature Selection and Decoder Training

Feature selection was performed on the training block by calculating the mean correlation coefficient (MCC)^17^ for each feature (computed as described in the previous paragraph). For each trial, data during the period from cue onset to 2 seconds after was extracted, normalized, and its temporal correlation to the average of all trials was calculated. Channels with an MCC above 0.45 were selected as inputs to the decoder.

### LSTM Decoder Architecture

The decoding was performed using a Long Short-Term Memory (LSTM) network. The architecture consisted of a sequence input layer, followed by an LSTM layer with 150 hidden units using a tanh activation function and a sigmoid gate. A dropout layer with a probability of 0.01 was incorporated to prevent overfitting, followed by a fully connected layer, and a softmax layer for classification. Grasp intention was decoded and updated at a frequency of 10 Hz.

### Functional Motor Task

We chose a functionally relevant task that was close to activities of daily living (ADLs) namely a simulated pouring task. The iSCI participant was instructed to grasp, tilt and hold a water-filled 700-gram glass bottle over a small mason jar, simulating a pouring action without any assistance. A successful trial was defined as when the iSCI participant grasped the bottle, maintained the grasp, initiated and held the bottle in a pouring position over the mason jar for at least 5 seconds without bottle/jar contact, placed the bottle down, and released the grasp. A failed trial was defined as dropping the bottle at any point during the task, not maintaining the bottle over the mason jar for at least 5 seconds, or the bottle contacting the mason jar during the ‘pour’. The iSCI participant was instructed to perform as many repetitions as possible within a predefined timeout period. Different timeout periods were implemented for completing the tasks but were usually 120, 180 or 240 seconds. The participants performed this task once per week and their baseline performance was recorded for up to 25 weeks.

### Cooperative Motor Control Task

To demonstrate the capacity for the avatar paradigm to enable shared motor control between two participants with SCI, the NMES device was secured to the forearm of the iSCI participant. Here, the participants were directly sitting across from each other, allowing for observation and communication throughout the task (Fig 3a). The LSTM-based decoder was a stable decoder generally developed as described above and described elsewhere^11^. It allowed for the decoding of both ‘open hand’ and ‘close hand’ signals from the iBCI participant. ‘Open hand’ signals triggered NMES electrodes on the dorsal side of the iSCI participant’s forearm, while ‘close hand’ signals triggered electrodes on the volar side. During this task, iBCI mediated transcutaneous spinal cord stimulation (tSCS) targeting C5-C6 dorsal spinal roots was delivered to the iSCI participant via a custom electrode patch, as described elsewhere^11^.

The participants were tasked with collaboratively performing the simulated pouring task. The same rules for successful and failed trials and timeout periods were applied as the unassisted version of the task.

To assess participant satisfaction and feedback, the participants were individually interviewed with a post-session questionnaire including the open-ended question: “What aspect of today’s experiments were you most satisfied with during today’s session?”. The question aimed to explore the aspects of the collaborative paradigm most valued by each participant, providing insights into the potential benefits of cooperative iBCI rehabilitation. Participant responses were video recorded and later transcribed.

### Transcutaneous spinal cord stimulation (tSCS)

The precise description of the stimulator, electrodes and dorsal root mapping is described elsewhere^11^. The iSCI participant was administered tSCS pulses paired with peripheral nerve stimulation in another ongoing experiment unrelated to the cooperative motor control task. tSCS and peripheral stimulation were temporally paired with precise timing for 3 bouts of 5 minutes, for a total of 15 minutes, once per week. The tSCS stimulation parameters were held at 250-300 mA. The peripheral nerve stimulation parameters were held at 200-250 mA. Both stimulation profiles consisted of sinusoidal waveforms with a pulse width of 0.5 ms and a frequency of 5 Hz. Additionally, 10-second-long stimulus trains were administered at the spinal cord to characterize the latency of the posterior root reflex, or at the periphery to characterize the latency of the evoked motor response. These experiments were carried out simultaneously as the baseline period for the unassisted pour task.

## Results

### Cortically Mediated Hand Movement in Human Avatar

The iBCI participant was able to successfully trigger the NMES and close the able-bodied participant’s hand within the allotted 15 second window via M1 decoding for all 21 object presentations in both cortically mediated trials, totaling 42/42 presentations. In all object presentations, a single, discrete, continuous grasp was maintained (Supplementary Fig. 1). The average intentional grasp length across both cortically mediated trials was 3.42 seconds with a standard deviation of 0.74 seconds (Supplementary Fig. 1c).

### Combining Spatial and Amplitude Encoding Paradigm for Discrimination

All stimulation paradigms yielded performance significantly above chance level (33.3%), as determined by an exact binomial test. Paradigm 8, which varied both number of electrodes simultaneously stimulated and stimulation amplitude, enabled the iBCI participant to accurately identify 93.3% (28/30) of presentations, more than any other paradigm (Table 1).

Paradigm 7, which varied by number of electrodes stimulated alone, was the second highest-performing paradigm at 27/30 (90%). We hypothesized that ramping of stimulation amplitude and/or frequency would help capturing anticipated differences in the rates at which objects of different compliance reached maximum force during grasping. While all 8 paradigms performed above chance, the three lowest-performing paradigms varied by ramping, or the rate at which the stimulation train reached its maximum value – at 0.5, 1.0 or 1.5 seconds into the 2 second stimulation train. Reported areas of sensory percepts from Paradigm 8 are included as a shaded drawing on the distal portion of the second digit (Fig 1c).

### Object Discrimination Task

In computer-mediated trials, grasping of the three objects produced distinct force profiles (Supplementary Fig 2a). Object discrimination accuracy during computer-mediated trials was 74.2% (89/120, *p* < 0.001; Supplementary Fig 2b). The iBCI participant was most accurate in identifying the soft foam ball (95%, 38/40, *p* < 0.001), but all three objects were identified individually at a rate significantly above chance (Dense foam: 75%, 30/40, *p* = 0.001; Baseball: 53%, 21/40, *p* = 0.010). 71.0% of errors (22/31) during computer-mediated trials occurred in discriminating between dense foam and baseball. Most errors (54.8%, 15/31) resulted from incorrectly guessing that the baseball was the dense foam ball. The dense foam ball was guessed most often (40.8% of responses, 45/120) and the baseball was guessed least often (21%, 26/120; Supplementary Fig. 2b).

In cortically mediated trials, the iBCI participant grasped the object during all 42 object presentations, indicating that successful M1 decoding was achieved even with brief, but simultaneous, S1 stimulation. Grasping resulted in distinct force profiles per object (Fig 2a) and subsequently distinct S1 stimulation profiles per object, as displayed by total current (amplitude X number of electrodes (Fig 2c and 2d). Discrimination accuracy in cortically mediated trials was 64.3% (27/42, *p* < 0.001; Fig 2b). The iBCI participant identified all three objects at the same rate, 64.3% (9/14, *p* = 0.017). Soft foam, dense foam, and baseball were guessed 30.1% (13/42), 45.2% (19/42) and 23.8% (10/42) of object presentations respectively. 60% (9/15) of errors were made in discriminating between soft foam and dense foam, and the remaining 40% (6/15) were made in discriminating between dense foam and baseball. In trials that resulted in incorrect guesses (Fig 2d) total current profiles for each object were less distinct and had greater overlap with the current profiles of the other objects as compared to trials that resulted in correct guesses (Fig 2c).

**Fig 2.**
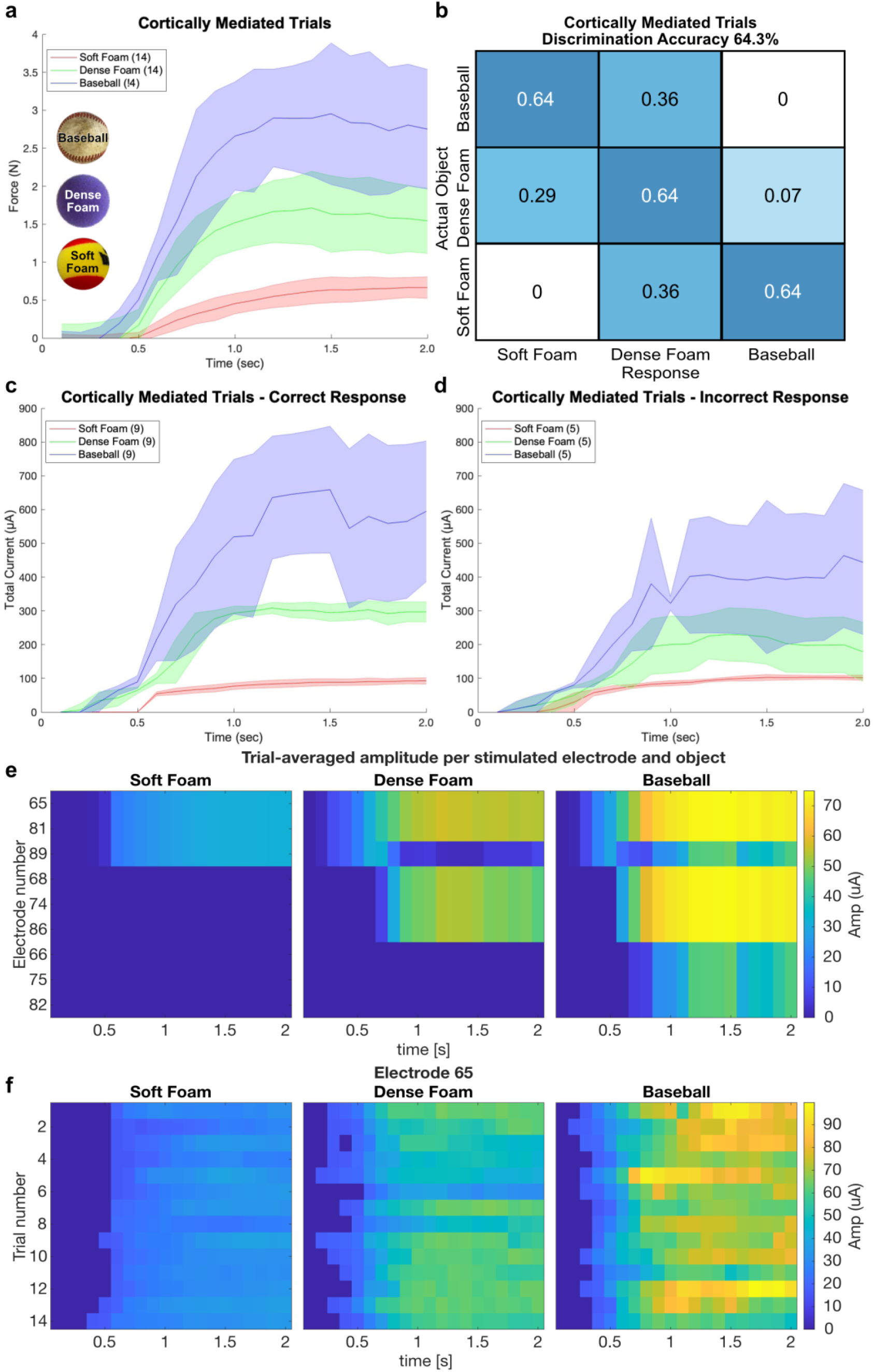
Results of cortically mediated object discrimination task. **a)** Mean (solid line) and standard deviation (shaded region) of force values recorded during the first 2 seconds of grasping in cortically mediated trials. Color coded by object identity: blue = baseball, green = dense foam ball, red = soft foam ball. **b)** Confusion matrix of the iBCI participant responses to object presentations during cortically mediated trials. 42 presentations over 2 trials, with an overall accuracy of 64.3%. **c)** Mean (solid line) and standard deviation (shaded region) of total current in μA of S1 stimulation during the first 2 seconds of grasping during cortically mediated trials that resulted in correct identification of the presented object. Total current calculated as a product of amplitude x number of electrodes. (Blue = Baseball, Green = Dense Foam, Red = Soft Foam). **d)** Mean (solid line) and standard deviation (shaded region) of total current in μA of S1 stimulation during the first 2 seconds of grasping during cortically mediated trials that resulted in incorrect identification of the presented object. (Blue = Baseball, Green = Dense Foam, Red = Soft Foam). **e)** Trial-averaged amplitude per stimulated electrode and object during the first two seconds after force onset. **f)** Stimulation delivered to S1 via electrode 65 (which belonged to the 3, 5, and 9 electrode subsets) per trial per object during the first two seconds after force onset

When asked to subjectively describe the stimulation delivered during object grasping, the iBCI participant reported that different objects felt ‘strong’ and ‘light.’ Although stimulation by different electrode numbers produced sensations in different finger areas (Fig. 1c), the iBCI participant said he mainly judged objects by “strong” or “light” intensity, not by location.

During both computer- and cortically mediated trials, the maximum total current during grasping of the three objects was significantly different. During the computer-mediated trials the maximum total currents registered were 80.97 µA, 304.22 µA, and 676.25 µA, for the soft foam, dense foam and baseball respectively (ANOVA, p-value: < 0.001). For the cortically mediated trials we observed 99.17 µA, 290.02 µA, and 659.98 µA as the maximum total currents for the three objects respectively (ANOVA, p-value: < 0.001). The target number of electrodes (3, 5, and 9 for soft foam, dense foam, and baseball, respectively) was reached for 85.8% (103/120, *p* < 0.001) of presentations during computer-mediated trials and 85.7% (35/42, *p* < 0.001) during cortically mediated trials.

### Human Avatar Paradigm Allows for Cooperative Movement During a Functional Task

Having demonstrated that the cortically interfaced avatar paradigm could restore both volitional motor control and discriminative touch via an able-bodied participant’s hand, we next sought to explore its potential for cooperative functional tasks between two participants with spinal cord injuries resulting in upper-limb deficits. We hypothesized that by modifying the avatar paradigm to enable the iBCI participant to control, via M1 decoding and NMES, the actions of the iSCI participant, we could create and explore the potential of a novel form of shared rehabilitation (Fig. 3a).

**Fig 3.**
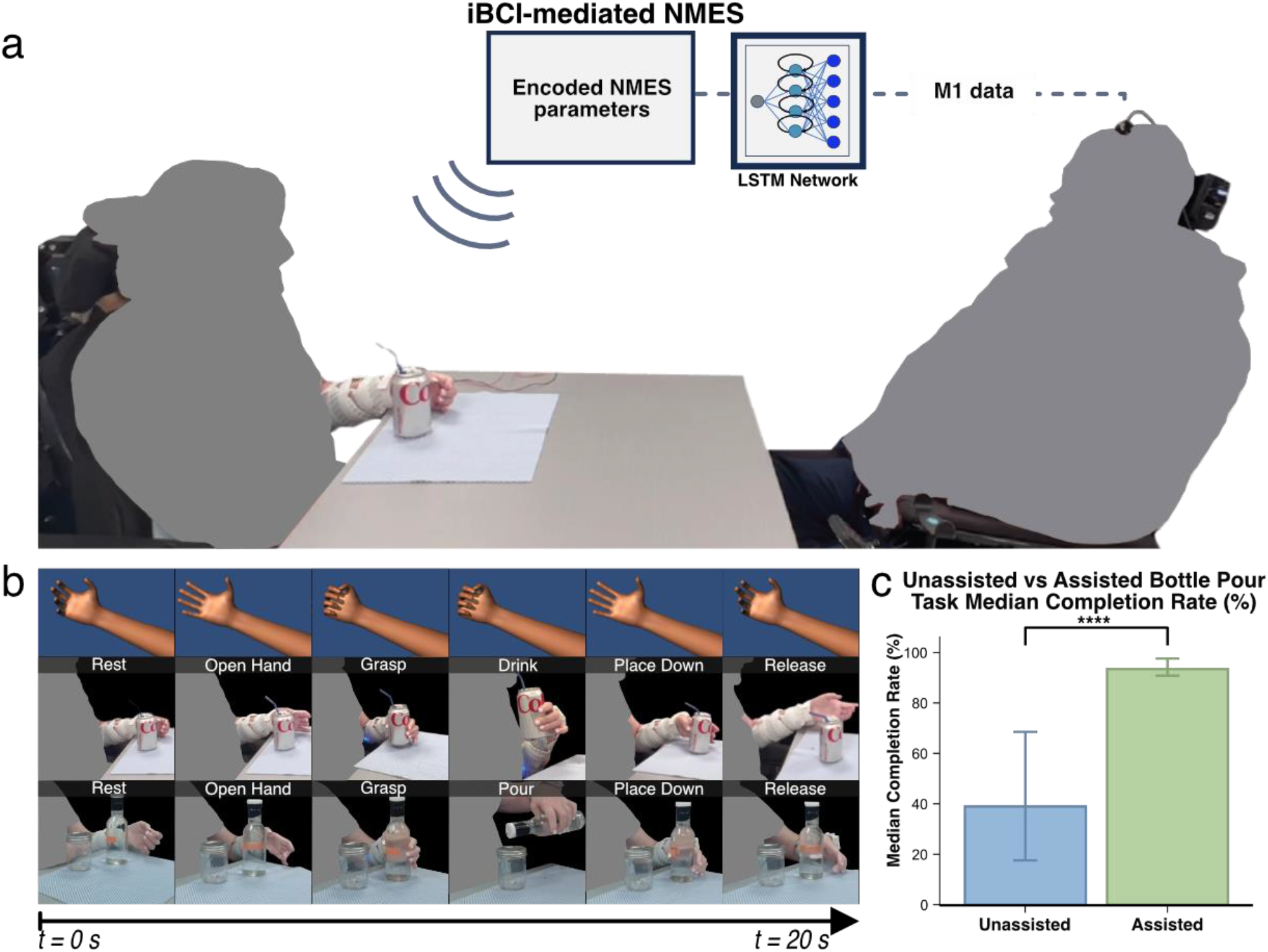
Cooperative rehabilitation experiment using a human avatar paradigm. a) Setup arrangement with the iSCI participant (left) across a table from the iBCI participant (right) **b)** Top row depicts computer animation stills of M1 decoder output from the iBCI participant during task. Middle and bottom rows depict corresponding grasping and releasing of the iSCI participant’s hand during two functional tasks, bottle self-feeding and pouring, respectively. **c)** Median completion rates of unassisted vs. tSCS+NMES assisted bottle pour task (39% vs 94% p = <0.0001 (Wilcoxon test))

Here, the iSCI participant verbally communicated her intended hand movements (“open”, “close”, and “hold”) to the iBCI participant. The iBCI participant then attempted the specified grasp movement (Fig. 3b, top), while a real-time decoder transmitted the intended movement wirelessly to activate NMES on the iSCI participant’s forearm, enabling her to self-feed (Fig. 3b, middle). Across several sessions, the participants collaborated to complete a water bottle grasping and pouring task (Fig. 3b, bottom) within a 120-second trial period, averaging approximately 20 seconds per repetition. iBCI-mediated tSCS+NMES assisted pouring trials had a 94% success rate (n=4 session) success rate compared to 39% success rate (n=23 sessions) without assistance (Fig. 3c).

Qualitative interviews revealed high satisfaction with the collaborative paradigm among both participants. The iBCI participant reported a sense of satisfaction when using the iBCI to contribute to the iSCI participant’s successful completion of the task. He stated that he found the paradigm engaging and that gave him a sense of purpose, stating “I was more satisfied [because] I was helping somebody in real life…rather than just a computer.” CTS05 also stated that she was satisfied with the effective communication between them, especially when utilizing streamlined commands, highlighting the importance of effective communication in achieving successful task completion. She acknowledged the iBCI participant’s crucial role in her success, stating “I wouldn’t have done that without you.” Overall, both participants valued the collaborative nature of the interaction and the shared sense of accomplishment.

## Discussion

In this work we developed and demonstrated a novel human avatar paradigm that can allow a person living with severe sensorimotor impairment to remotely interact with objects, through another person, using interhuman neural bypass technology. This approached linked decoded movement intention from an implanted bidirectional brain-computer interface in one person to neuromuscular activation wirelessly in another person. This paradigm also allowed two participants with tetraplegia to engage in cooperative rehabilitation, demonstrating increased success in a motor task with a real-world object.

The novel cortically interfaced human avatar paradigm provided several advantages. First, the avatar allowed us to avoid confounding effects of any residual sensory input in the iBCI participant as the effectiveness of S1 in object discrimination was evaluated. Consequently, the only intentional feedback provided for the discrimination of objects was via S1 ICMS. Additionally, using an avatar avoided confounding factors such as decreased strength and endurance of the iBCI participant’s grasp due to atrophy since his injury. In summary, the avatar approach allowed for a controlled experimental environment but could allow for robust motor-sensory integration even in patients in whom direct muscle stimulation may be severely limited. The selection of a stimulation paradigm followed from testing of eight distinct options, such as varying stimulation amplitude, frequency, electrode number, and combinations therein. Notably, varying both number of electrodes stimulated, and stimulation amplitude simultaneously resulted in the greatest performance in discrimination of three stimulation intensities. Variation in number of electrodes allows for providing a greater dynamic range of perception intensities without exceeding safety limits for current density while delivering ICMS. In addition, the participant’s reported differences in percept area on their index finger highlight the distinct somatotopic areas elicited by distinct subsets of electrodes. Although not the top-performing paradigm, varying number of electrodes alone also resulted in high discrimination accuracy (90%). This result appears to contradict evidence from non-human primate studies, in which variation in number of electrodes only has a marginal effect on discriminability of stimulation trains^18^. The benefit of multielectrode stimulation to discriminability may also be augmented in humans by a greater degree of cortical plasticity and adaptation to chronic microstimulation, although these parameters have not been directly compared to non-human primates^19^.

Object discrimination with performance significantly above chance was achieved in cortically mediated trials. Analysis of the total current profiles of correct vs. incorrect trials revealed less distinct profiles in incorrect trials. This overlap was also reflected in the force profiles recorded from sensors on the iBCI-participant’s fingers. We conclude it is likely that a proportion of incorrect guesses were a result of inadequate or oblique contact between the object and the force sensors. Despite these limitations, the distinct stimulation profiles per object and the high rates of reaching the target number of electrodes indicate that the previously stated goals which motivated the design of the encoding algorithm were largely met. In future experiments we expect performance to improve with thinner force sensors which will optimize contact and result in more consistently distinct force profiles, and therefore total current profiles per object.

Additionally, we demonstrated a novel paradigm of iBCI-mediated cooperative rehabilitation wherein two individuals with paralysis cooperated with each other while receiving activity-based stimulation, a largely unexplored approach. Traditionally physical therapy has been focused on the individual performing the exercise with some motivation and help from their therapist. This setting has its limitation in terms of limited time available with the therapist and the lack of motivators when the patient is at home. Recent studies have shown that having a paradigm where patients work with a family member^20^ or even other participants^21^ while performing tasks appear to increase motivation among participants. Studies also show that such interaction need to have both a visual as well as a physical component to the interaction to be most effective^22^. Our cooperative approach supports this, where we found that this paradigm did promote positive interactions, increased motivation, and a sense of accomplishment between the SCI participants, thereby benefiting both.

The potential benefits of bidirectional iBCI technology, in addition to the avatar paradigm, should be explored in the context of nervous system pathologies such as upper-limb stroke, motor neuron disease, and peripheral nerve damage. About 85% of stroke victims suffer from residual upper limb disability^23^. Recovery of motor function after stroke relies on neuroplastic processes - several forms of iBCI, most commonly non-invasive electroencephalography, have been shown to promote long-term clinical and neurophysiological change in stroke patients^24^. Motor imagery (MI), defined broadly as the mental rehearsal of intended movements, has recently been shown to be beneficial as an adjunct to traditional motor therapy in upper-limb stroke patients^25^. The therapeutic value of MI likely lies in the activation of similar or overlapping neural networks as the ones which are damaged or inhibited after stroke. During MI, patients are asked to imagine ‘external’ or ‘internal’ images – the intended motion either from a 3rd person perspective, or in the 1st person. The bidirectional iBCI paradigm described in our experiments presents an opportunity to record and decode movement intention from stroke patients, and to provide specific, easily modulated sensory feedback via S1 stimulation. The demonstrated shared motor task between two SCI participants highlights the potential therapeutic applications of the human avatar paradigm.

Careful selection of an encoding algorithm for S1 ICMS allowed for robust performance in the object discrimination task. In object discrimination tasks utilizing spinal cord stimulation, grasp force has been encoded to stimulation amplitude both linearly and exponentially, allowing for discrimination of objects by size and compliance^26^. A similar linear encoding scheme between grasp force and stimulation amplitude has been applied to peripheral nerve stimulation^27^. Other groups have encoded both proprioceptive and force information collected from a myoelectric prosthetic hand to peripheral nerve stimulation frequency^28^.

ICMS has been shown to evoke somatotopically specific percepts in the hands after surgical electrode implantation in S1 – these perceptions also scale linearly with stimulation amplitude. This known linear relationship between stimulation amplitude and percept intensity in both spinal cord and intracranial studies informed the selection of a split-linear encoding algorithm^9,29^. In that study, electrodes corresponding to distinct hand regions were stimulated according to where the virtual prosthetic limb contacted the virtual objects, which were a uniform cylinder and two objects composed of four cylindrical segments of descending or ascending diameter. S1 ICMS with multiple electrodes has been shown to elicit a percept which approximates the sum of the receptive fields of each individual electrode, in addition to providing a larger range of perceived intensities and more discrete levels when compared to single electrode stimulation (19.5 vs 11). Interestingly, amplitude intensity functions still follow a linear relationship with multiple electrode stimulation, as they do with single electrodes^19^. It is unknown if varying both electrode number and amplitude provides an even greater range of intensities and discrete levels or follows a linear relationship with perceived intensity, although the results reported here suggest their combination improves overall discrimination. Until the present study, a combined encoding algorithm which exploits both varying amplitude and number of electrodes to encode stimulus intensity has yet to be utilized in an object discrimination task.

Our findings demonstrate that simultaneous M1 decoding and S1 stimulation through a bidirectional iBCI can enable a person with SCI to achieve voluntary grasping and object discrimination via another individual’s hand. This paradigm supports both remote sensorimotor interaction—with broad implications—and cooperative rehabilitation with direct clinical relevance. The sensorimotor human avatar provides robust visual and tactile feedback to individuals with upper-limb motor deficits, highlighting its potential as a novel therapeutic approach. By combining a bidirectional iBCI with targeted neurostimulation, we enabled a participant with profound sensorimotor loss to establish a unique connection with a partner through shared task. Crucially, this approach not only restores aspects of sensorimotor function but also fosters interpersonal connection, allowing individuals with paralysis to re-experience agency, touch, and collaborative action through another person. Beyond rehabilitation, this paradigm opens new avenues for exploring how neural interfaces can extend human capability, enrich shared experiences, and deepen our connections.

## Data Availability

All data produced in the present study are available upon reasonable request to the authors

## Notes

### Competing Interest Statement

Disclosures: E. Cater: None. S. Chandrasekaran: None. S Wandelt: None. A. Jangam: None. Z. Elias: None. E Ibroci: None. C. Maffei: None. S Bickel: None. N Ben-Shalom: None. A Stein: None. A Mehta: None. C Bouton: Financial interests in Neuvotion and Sanguistat.

### Clinical Trial

NCT03680872 and NCT04755699

### Funding Statement

This study was funded by New York State Department of Health Spinal Cord Injury Research Board (Contract #C37718GG) and the Feinstein Institutes for Medical Research at Northwell Health with additional support from Blackrock Neurotech and Good Shepherd Rehabilitation Network.

### Author Declarations

The study was conducted under a Food and Drug Administration Investigational Device Exemption #G170200. IRB of Northwell Health gave ethical approval for this work (IRB protocols #17-0840 (NCT03680872) and #20-0425 (NCT04755699)), in accordance with the Declaration of Helsinki.

## References

1. Hochberg, L. R. et al. Reach and grasp by people with tetraplegia using a neurally controlled robotic arm. Nature 485, 372–5 (2012).

2. Bouton, C. E. et al. Restoring cortical control of functional movement in a human with quadriplegia. Nature 533, 247–250 (2016).

3. Ajiboye, A. B. et al. Restoration of reaching and grasping in a person with tetraplegia through brain-controlled muscle stimulation: a proof-of-concept demonstration. Lancet Lond. Engl. 389, 1821–1830 (2017).

4. Sharma, G. et al. Using an Artificial Neural Bypass to Restore Cortical Control of Rhythmic Movements in a Human with Quadriplegia. Sci. Rep. 6, 33807 (2016).

5. Collinger, J. L. et al. High-performance neuroprosthetic control by an individual with tetraplegia. The Lancet 381, 557–564 (2013).

6. Willsey, M. S. et al. A high-performance brain–computer interface for finger decoding and quadcopter game control in an individual with paralysis. Nat. Med. 31, 96–104 (2025).

7. Armenta Salas, M. et al. Proprioceptive and cutaneous sensations in humans elicited by intracortical microstimulation. eLife 7, e32904 (2018).

8. Fifer, M. S. et al. Intracortical Microstimulation Elicits Human Fingertip Sensations. medRxiv 2020.05.29.20117374 (2020) doi:10.1101/2020.05.29.20117374.

9. Flesher, S. N. et al. Intracortical microstimulation of human somatosensory cortex. Sci. Transl. Med. 8, (2016).

10. Flesher, S. N. et al. A brain-computer interface that evokes tactile sensations improves robotic arm control. Science 372, 831–836 (2021).

11. Chandrasekaran, Santosh et al. Restoring Cortically Mediated Movement and Sensation in Complete Tetraplegia. 2025.08.19.25330198 Preprint at 10.1101/2025.08.19.25330198 (2025).

12. Metzger, S. L. et al. A high-performance neuroprosthesis for speech decoding and avatar control. Nature 620, 1037–1046 (2023).

13. Mann, J. The medical avatar and its role in neurorehabilitation and neuroplasticity: A review. NeuroRehabilitation 46, 467–482 (2020).

14. Eden, J. et al. Principles of human movement augmentation and the challenges in making it a reality. Nat. Commun. 13, 1345 (2022).

15. Willett, F. R. et al. A high-performance speech neuroprosthesis. Nature 620, 1031–1036 (2023).

16. Rao, R. P. N. et al. A Direct Brain-to-Brain Interface in Humans. PLoS ONE 9, e111332 (2014).

17. Bouton, C. et al. Decoding Neural Activity in Sulcal and White Matter Areas of the Brain to Accurately Predict Individual Finger Movement and Tactile Stimuli of the Human Hand. Front. Neurosci. 15, 699631 (2021).

18. Kim, S., Callier, T., Tabot, G. A., Tenore, F. V. & Bensmaia, S. J. Sensitivity to microstimulation of somatosensory cortex distributed over multiple electrodes. Front. Syst. Neurosci. 9, (2015).

19. Huang, Y.-Z., Chen, R.-S., Rothwell, J. C. & Wen, H.-Y. The after-effect of human theta burst stimulation is NMDA receptor dependent. Clin. Neurophysiol. 118, 1028–1032 (2007).

20. Baur, K. et al. Robot-Supported Multiplayer Rehabilitation: Feasibility Study of Haptically Linked Patient-Spouse Training. in 2018 IEEE/RSJ International Conference on Intelligent Robots and Systems (IROS) 4679–4684 (2018). doi:10.1109/IROS.2018.8593769.

21. Thielbar, K. O. et al. Home-based Upper Extremity Stroke Therapy Using a Multiuser Virtual Reality Environment: A Randomized Trial. Arch. Phys. Med. Rehabil. 101, 196–203 (2020).

22. Küçüktabak, E. B., Kim, S. J., Wen, Y., Lynch, K. & Pons, J. L. Human-machine-human interaction in motor control and rehabilitation: a review. J. NeuroEngineering Rehabil. 18, 183 (2021).

23. Nichols-Larsen, D. S., Clark, P. C., Zeringue, A., Greenspan, A. & Blanton, S. Factors Influencing Stroke Survivors’ Quality of Life During Subacute Recovery. Stroke 36, 1480–1484 (2005).

24. Carvalho, R., Dias, N. & Cerqueira, J. J. Brain-machine interface of upper limb recovery in stroke patients rehabilitation: A systematic review. Physiother. Res. Int. J. Res. Clin. Phys. Ther. 24, e1764 (2019).

25. Machado, T. C., Carregosa, A. A., Santos, M. S., Ribeiro, N. M. D. S. & Melo, A. Efficacy of motor imagery additional to motor-based therapy in the recovery of motor function of the upper limb in post-stroke individuals: a systematic review. Top. Stroke Rehabil. 26, 548–553 (2019).

26. Nanivadekar, A. C. et al. Closed-loop stimulation of lateral cervical spinal cord in upper-limb amputees to enable sensory discrimination: a case study. Sci. Rep. 12, 17002 (2022).

27. Raspopovic, S. et al. Restoring Natural Sensory Feedback in Real-Time Bidirectional Hand Prostheses. Sci. Transl. Med. 6, (2014).

28. Horch, K., Meek, S., Taylor, T. G. & Hutchinson, D. T. Object discrimination with an artificial hand using electrical stimulation of peripheral tactile and proprioceptive pathways with intrafascicular electrodes. IEEE Trans. Neural Syst. Rehabil. Eng. Publ. IEEE Eng. Med. Biol. Soc. 19, 483–489 (2011).

29. Chandrasekaran, S. et al. Correction: Sensory restoration by epidural stimulation of the lateral spinal cord in upper-limb amputees. eLife 10, e72438 (2021).

