## Supplemental Figures for "Cortically Interfaced Human Avatar Enables Remote Volitional Grasp and Shared Discriminative Touch"

### Supplementary Figures

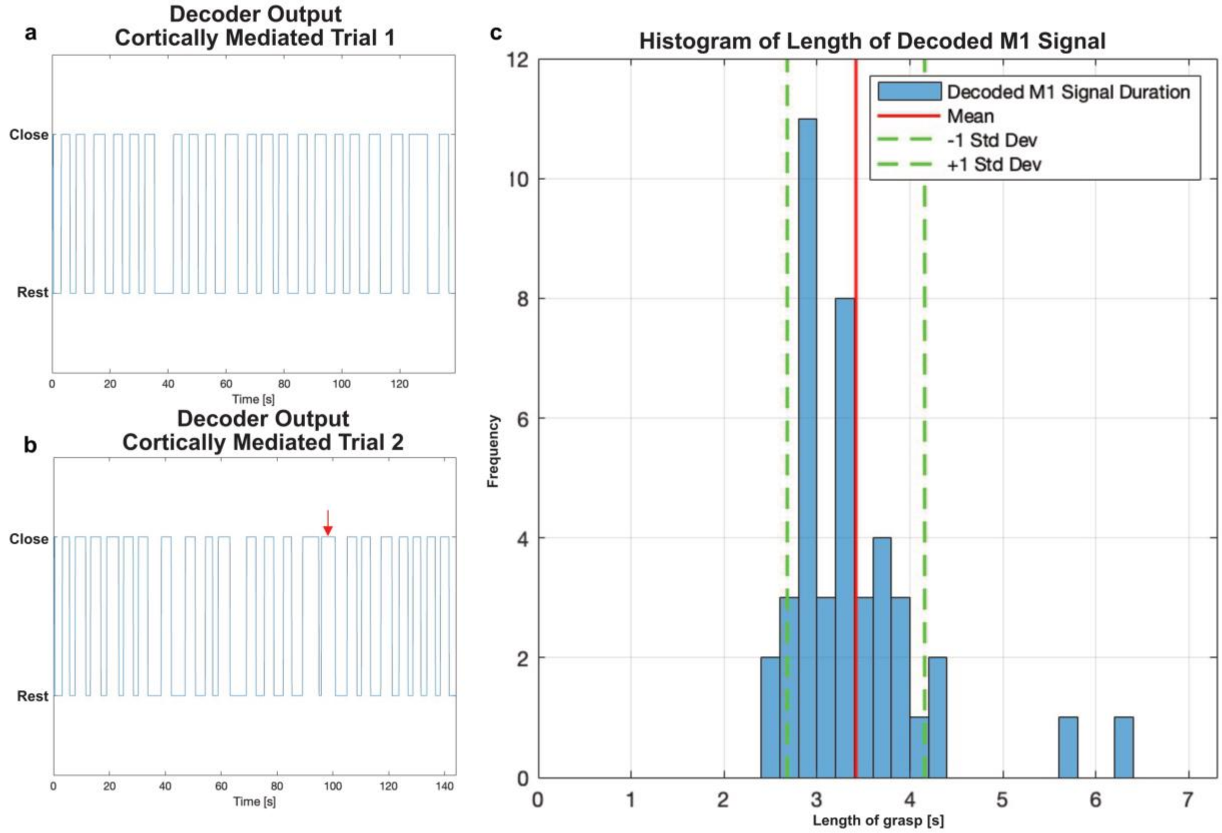


***Supplementary Fig 1. Results of M1 decoding a)*** *Decoder output during the first of two cortically mediated trials.* ***b)*** *Decoder output during the second cortically mediated trial. Red arrow indicates the only unintentional grasp between object presentations.* ***c)*** *Histogram of the length of grasps. Grasps were sustained for a mean of 3.43 seconds, Stdev = 0.74.*

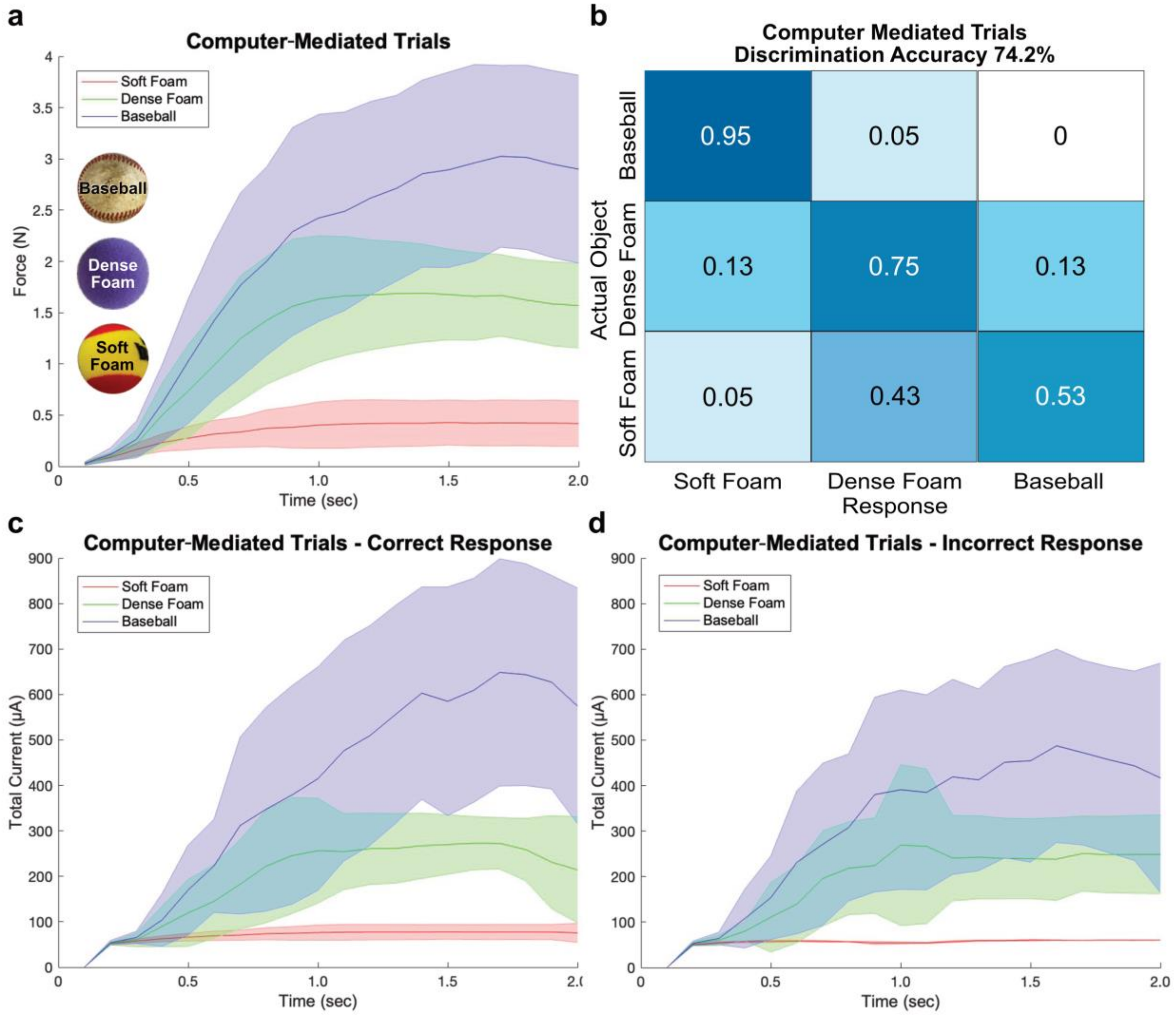


***Supplementary Fig 2. Results of computer-mediated object discrimination task a)*** *Mean (solid line) and standard deviation (shaded region) of force values recorded during the first 2 seconds of grasping in computer-mediated trials. Color coded by object identity: blue = baseball, green = dense foam ball, red = soft foam ball.* ***b)*** *Confusion matrix of participant responses to object presentations during computer-mediated trials. 120 presentations over 4 trials, with an overall accuracy of 74.2%.* ***c)*** *Mean (solid line) and standard deviation (shaded region) of total current in μA of S1 stimulation during the first 2 seconds of grasping during computer-mediated trials that resulted in correct identification of the presented object. Total current calculated as a product of amplitude x number of electrodes. (Blue = Baseball, Green = Dense Foam, Red = Soft Foam).* ***d)*** *Mean (solid line) and standard deviation (shaded region) of total current in μA of S1 stimulation during the first 2 seconds of grasping during computer-mediated trials that resulted in incorrect identification of the presented object. (Blue = Baseball, Green = Dense Foam, Red = Soft Foam).*
